# Influence of Novel Coronavirus COVID – 19 and HIV: A Scoping Review of Hospital Course and Symptomatology

**DOI:** 10.1101/2021.07.25.21260967

**Authors:** Mona Sheikh, Shavy Nagpal, Madiha Zaidi, Rupalakshmi Vijayan, Wanessa Matos, Neguemadji Nagardig Ngaba, Lordstrong Akano, Samia Jahan, Shazia Q. Shah, Camille Celeste Go, Sindhu Thevuthasan, George Michel

## Abstract

**Background:** An outbreak of novel coronavirus (SARS- CoV-2) was observed on December 2019 in Wuhan, China which led to a global pandemic declared in March 2020. As a consequence, it imposed delirious consequences in patients with underlying co – morbid conditions that make them immunocompromised. The purpose of this paper is to provide an in - depth review of influence of COVID – 19 in patients with underlying HIV in terms of mortality and hospitalization.

Authors also aim to provide a thorough risk analysis of hospitalization, ICU admission and mortality of PLWH and COVID-19. The secondary objective was to analyze the CD4+ count variations and outcome of COVID - 19 and to correlate if ART provided a protective role. Authors also aim to provide an evaluation of typical clinical presentation of COVID-19 in PLWH. ART is found to show activity against SARS-CoV-2 in vitro, and there is some similarity in the structure of HIV-1 gp41 and S2 proteins of SARS-CoV since they both belong to +ssRNA type.

**Methods:** We conducted a literature review using search engines namely, Cochrane, PubMed and Google Scholar. The following keywords were targeted: “COVID-19,” “SARS-CoV-2,” and “HIV.” We included case reports, case series, and cohort (retrospective and prospective) studies. We excluded clinical trials and review articles. We came across 23 articles that met the inclusion criteria. PRISMA guidelines were followed for study acquisition (Fig. 9).

**Results:** From the 23 studies, we found a total of 651 PLWH with confirmed COVID-19 (549, 91, and 11 in cohorts, case series, and case reports, respectively). The overall risk of hospital admission from pooled data of the 23 reviewed articles was 69.13% (450/651), ICU admission was 12.90% (84/651) in total infected patients, and 18.67% (84/450) among hospitalized patients. The overall case fatality rate from the 23 reviewed articles was 11.21 (73/651).A weak positive correlation was found between CD4+ counts and hospital admissions in case series and case reports, while the weak negative correlation was found in cohorts. For mortality, there was a negative weak association in the cohorts and in case series, while a weak positive was seen in case reports (Fig.7). We assessed the presenting symptoms of PLWH with COVID-19, and our review demonstrated this group does not greatly differ from the rest of the population, as their common presenting symptoms were cough, fever, and SOB.

**Conclusion:** Our results indicated that there was a high rate of hospitalization, ICU admission, and mortality among patients living with HIV and COVID-19. PLWH needs to be noted as a high-risk group for COVID-19 complications and severity. We recommend that PLWH be closely monitored by their physicians and strictly adhere to antiretroviral therapy and standard universal COVID-19 precautions.

## Introduction

SARS-CoV-2 is a member of β-CoVs (Fig. 8) and is a global concern at present. What interests Authors were the overlapping similarities between the Human Immunodeficiency Virus (HIV) and SARS-COV-2 virus in terms of same RNA single-stranded family and protein structural motifs of HIV-1 gp41 and SARS-CoV-S2 [9]. Having said that, it was proposed in clinical trials to implement ART as a treatment modality for COVID – 19. However, it is not recommended by the Treatment Guidelines Panel for COVID-19 yet [10].

The prognosis of COVID-19 in common comorbidities such as HTN, DM and CVD is well known but its prognosis or outcome of patients living with HIV is not well understood so far (Ssentongo et al, 2020) [11]. This review article focuses on the people living with HIV which are confirmed COVID-19 positive cases. The main focus of our review is to assess the risk of hospitalization, intensive care unit (ICU) admission, invasive mechanical ventilation (IMV) requirement and mortality in persons living with HIV (PLWH) who were COVID-19 positive. The secondary objective is to assess whether any correlation exists between CD4+ count and COVID-19 in PLWH with the third objective to evaluate the typical clinical presentation of COVID-19 in such patients.

## Methods

### Literature search

A literature search was conducted with PubMed, Cochrane, and Google scholars using the following keywords: “COVID-19”, “HIV,” “SARS-CoV-2”, and “AIDS” from January 2020 until August 31st, 2020. Only papers in English language were considered. PRISMA guidelines were followed (Fig. 9).

### Inclusion & Exclusion criteria and study selection

The case reports, case series, retrospective cohort studies and prospective cohort studies published between January 1, 2020, and August 31, 2020 with data of PLWH and confirmed COVID-19 by RT-PCR were included in our review. Review articles, clinical trials, and preprint articles were excluded. Articles that did not have patient data and those limited to specific comorbidities and organ dysfunctions were also excluded to avoid selection bias.

We narrowed it down to 48 articles that were screened based on the relevancy of the titles and abstracts. We excluded 32 articles, of which 20 did not meet the inclusion criteria, 2 required access, and 3 were not peer-reviewed. We included 23 studies in this paper. Selected articles were independently reviewed by two authors. All disagreements were resolved with a discussion between the two authors or with input from a third independent reviewer and mutually agreed upon by the authors.

### Data Extraction

The authors designed a predefined data extraction list to be used to extract the required data from the included studies. The following data were extracted: First author’s name, publication date, study design, country, the total number of PLWH with confirmed COVID-19 by RT-PCR or antigen test, age, gender, comorbidities, ART, CD4+ cell counts, viral load, number of hospitalized patients (age, gender), duration of hospitalization, number admitted to ICU, a number that needed IMV and number of deaths (age, gender, comorbidity, CD4+ counts, ART list).

### Data collection and Analysis

The data was collected on excel sheets and Google documents. We ran a qualitative review based on our primary and secondary outcomes. The studies used have been listed in Table 1-3. The data were tabulated using Microsoft Excel.

**Table 1.** Outcome of PLWH with COVID-19: Cohort Studies.

| Author/Study type | Location | Total no. infect. pts | Total no. hosp. pts | Rate of hosp. | Total no. ICU pt. | ICU rate (total pt., hosp.) | Total no. IMV pt. | Rate of IMV (total pt., hosp., ICU) |
| --- | --- | --- | --- | --- | --- | --- | --- | --- |
| Viscarra et al/<br>Prospective | Spain | 35 | 28 | 80% | 6 | 21.4%, 17.1% | 5 | 14.3%, 17.9% |
| Del Amo et al/Prospective | Spain | 236 | 151 | 64% | 15 | 9.9%, 6.4% | NA | N/A |
| Ho et al/<br>Retrospective | USA | 93 | 72 | 77.40% | 19 | 20.4%, 26.4% | 15 | 16.1%, 20.8% |
| Sigel et al /<br>Retrospective | USA | 88 | 88 | 100% | 18 | 20.5%, 205% | 18 | 20.5%, 20.5% |
| Meyerowitz et al /<br>Retrospective | USA | 36 | 21 | 58% | 6 | 28.6%, 16.6% | 4 | 11.1%, 19.1% |
| Gervasoni et al /<br>Retrospective | Italy | 28 | 13 | 50% | 2 | 15.4%, 7.1% | 2 | 4.3%, 15.4% |
| Harter et al / Retrospective | Germany | 33 | 14 | 42% | 6 | 42.9%, 18.2% | 4 | 12.1%, 28.6% |
| Total |  | 549 | 387 | 70.49% | 72 | 13.11%, 18.61% | 48 | 8.74%, 12.70%, 66.67% |

**Table 2.** Outcome of PLWH with COVID-19: Case Series.

| Author/Study type | Location | Total no. infect pts. | Total no. hosp. pts | Rate of hosp | Total no. ICU pts | ICU rate (total pt., hosp.) | Total no. IMV pts. | Rate of IMV (total pt., hosp., ICU) |
| --- | --- | --- | --- | --- | --- | --- | --- | --- |
| Blanco et al / Case Series | Spain | 5 | 5 | 100% | 2 | 40%, 40% | 1 | 20%, 20% |
| Childs et al / Case Series | USA | 18 | 18 | 100% | 5 | 27.7%, 27.7% | 5 | 27.78%, 27.8% |
| Ridgeway et al / Case Series | USA | 5 | 5 | 100% | 1 | 20%, 20% | 0 | 0 |
| Byrd et al / Case Series | USA | 27 | 9 | 33.30% | 0 | 0 | 0 | 0 |
| Altuntas Aydin et al / Case Series | Turkey | 4 | 4 | 100% | 1 | 25%, 25% | 1 | 25%, 25% |
| Marimuthu et al / Case Series | India | 6 | 6 | 100% | 0 | 0 | 0 | 0 |
| Calza et al / Case Series | Italy | 26 | 5 | 19.20% | 0 | 0 | 0 | 0 |
| Total |  | 91 | 52 | 57.14% | 9 | 9.89%, 17.31% | 7 | 7.69%, 13.46%, 77.78% |

**Table 3.**
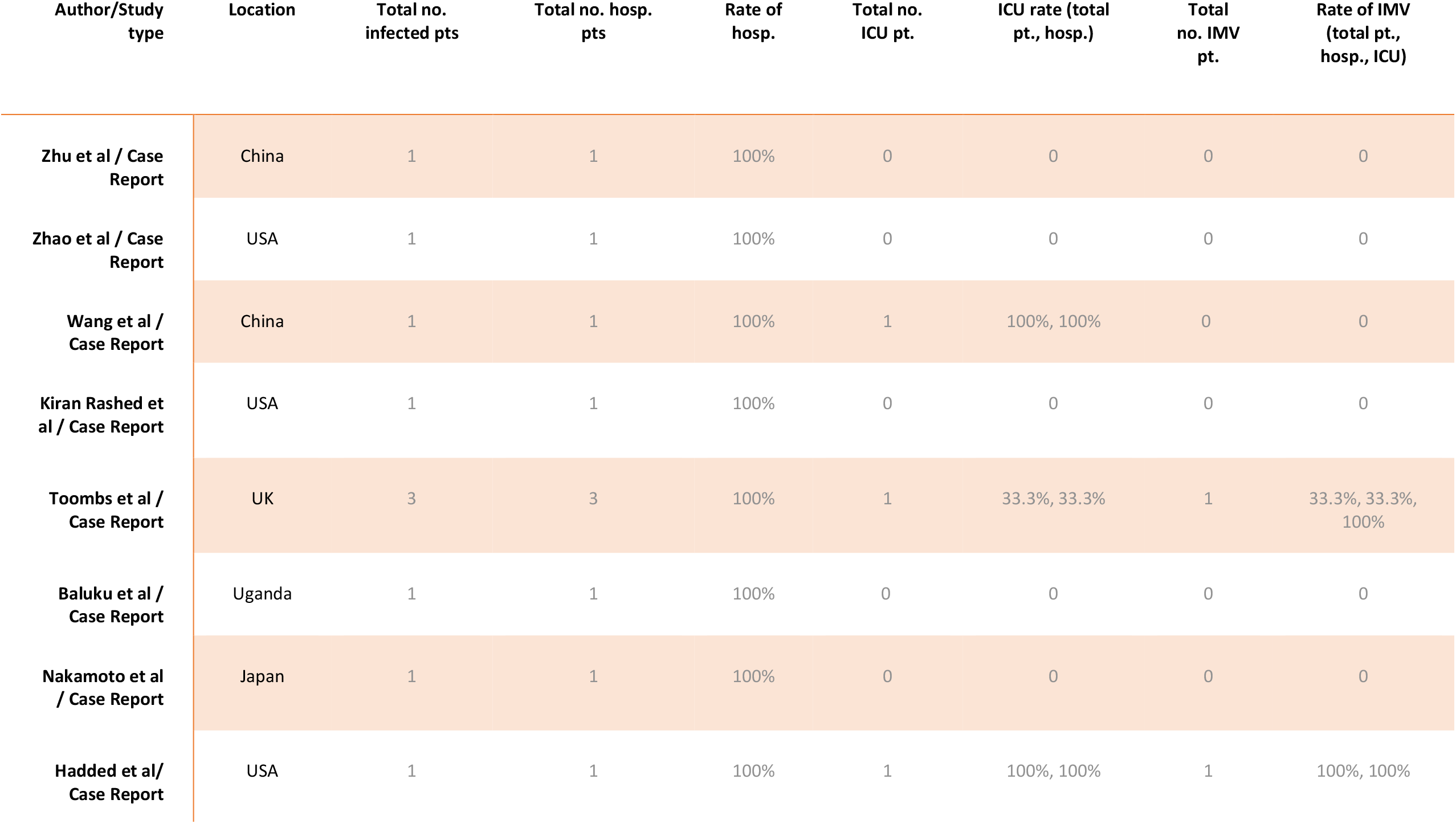
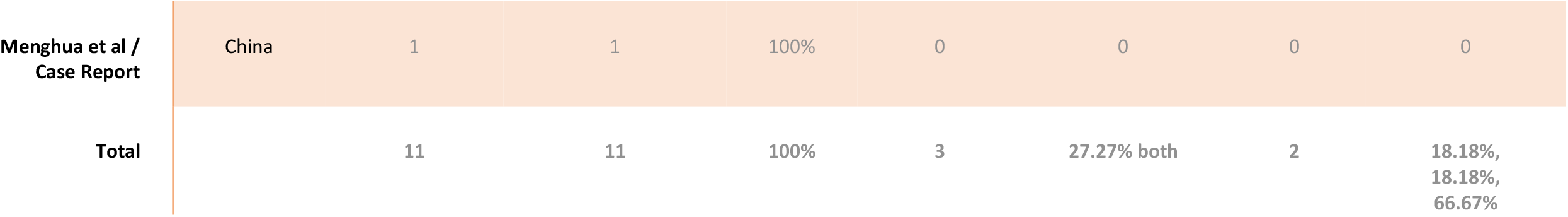
Outcome of PLWH with COVID-19: Case Reports.

The Pearson correlation coefficient was used to assess the correlation between CD4+ counts and hospitalization and mortality, and the association of COVID-19 infection with hospital admission. A P-value <0.05 was considered significant and was calculated by using R factor, Z score and standard deviation.

### Ethical approval and funding

We did not seek IRB approval, as our study is based on data from published articles and patients were not directly involved. No funding was obtained for this review.

## Results

This review included a total of 23 articles after excluding those that did not meet the inclusion criteria (20), not peer reviewed (3), and required access (2). The 23 articles were 7 cohort studies, 7 case series, and 9 case reports; (Fig. 10) and they were from different countries: USA (9), China (4), Spain (3), Italy (2), the UK (1), Germany (1), Turkey (1), India (1), Japan (1) and Uganda (1) (Fig. 11)

Our review was categorized into three groups based on the similarity of study type: which are cohort group (Ch), case series group (CS), and case report group (CR). The number of PLWH and confirmed COVID-19 were 549 from cohort studies (Ch), 91 from case series (CS), and 11 from case reports (CR). The mean age (in years) were 54 in Ch, 45 in CS, and 36 in CR. The gender distribution in the Ch was 440 male, 106 female, and 3 transgender; in CS was 62 male, 25 female, and 3 transgender; and in CR was 8 males and 3 females. [Tables 1-3]

### Risk of hospitalization of PLWH + confirmed COVID-19

The total number of hospitalized patients were 387 in Ch, 52 in CS, and 11 in CR. The risk of hospitalization in these 3 groups were 70.49% (387/549) in Ch, 57.14% (52/91) in CS, and 100% in CR. [Table 1-3] The overall risk of hospital admission from pooled data of the 23 reviewed articles was 69.13% (450/651).

### Risk of ICU admission among hospitalized patients

The number of patients that needed critical care were 72 in Ch, 9 in CS, and 3 in CR. The calculated risk among hospitalized patients was 18.61% (72/387) in Ch, 17.31% (9/52), and 27.27% (3/11) in CR. [Table 1-3] The overall risk of ICU admission from pooled data of the 23 reviewed articles was 12.90% (84/651) in total infected patients and 18.67% (84/450) among hospitalized patients.

### The risk of IMV requirement in hospitalized and ICU patients

Of those patients who were admitted to the ICU: 48 in Ch, 9 in CS, and 2 patients in CR needed invasive mechanical ventilation (IMV). The risk among hospitalized and ICU patients was (12.7%, 66.67%) in Ch, (13.46%, 77.78%) in CS, and (18.18%, 66.67%) in CR. [Table 1-3]

### The mortality rate of PLHW + confirmed COVID-19

The total number of deaths in the three categories was 66 in Ch, 6 in CS, and 1 CR, respectively. The case fatality rate was 12.02% (66/549) in Ch, 6.59% (6/91) in CS, and 9.09% (1/11) in CR [Tables 1-3]. The overall case fatality rate from the 23 reviewed articles was 11.21 (73/651)

### Correlation of CD+ counts with hospitalization and mortality

The Pearson Correlation coefficient was used to assess the association of CD4+ counts with hospitalization and mortality in PLWH+COVID-19. A weak positive correlation was found between CD4+ counts and hospital admissions in case series (r = 0.2622, p-value = 0.089403) (Fig. 1) and case reports (r = 0.2269, p-value = 0.5284) (Fig. 2), while the weak negative correlation was found in cohorts (r = −0.114, p-value = 0.043866) (Fig. 3). For the mortality correlation, there was a negative weak association in the cohorts (r = −0.1218, p-value = 0.031) (Fig. 4) and in case series (r = −0.011, p-value = 0.986) (Fig. 5), while a weak positive was seen in case reports (r = 0.0667) (Fig. 6).

**Figure 1.**
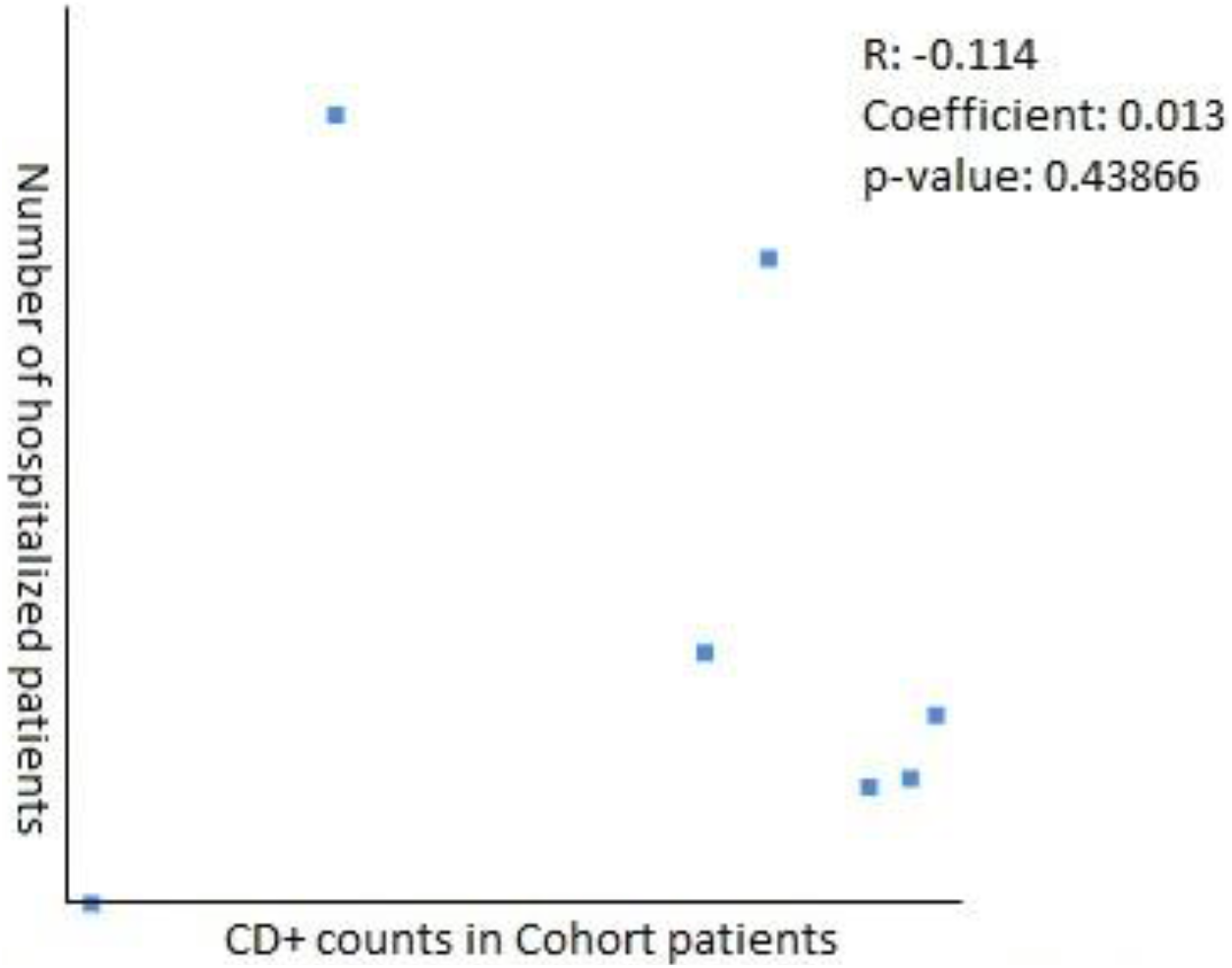
correlation of CD4+ counts & hospitalization in cohorts patients

**Figure 2.**
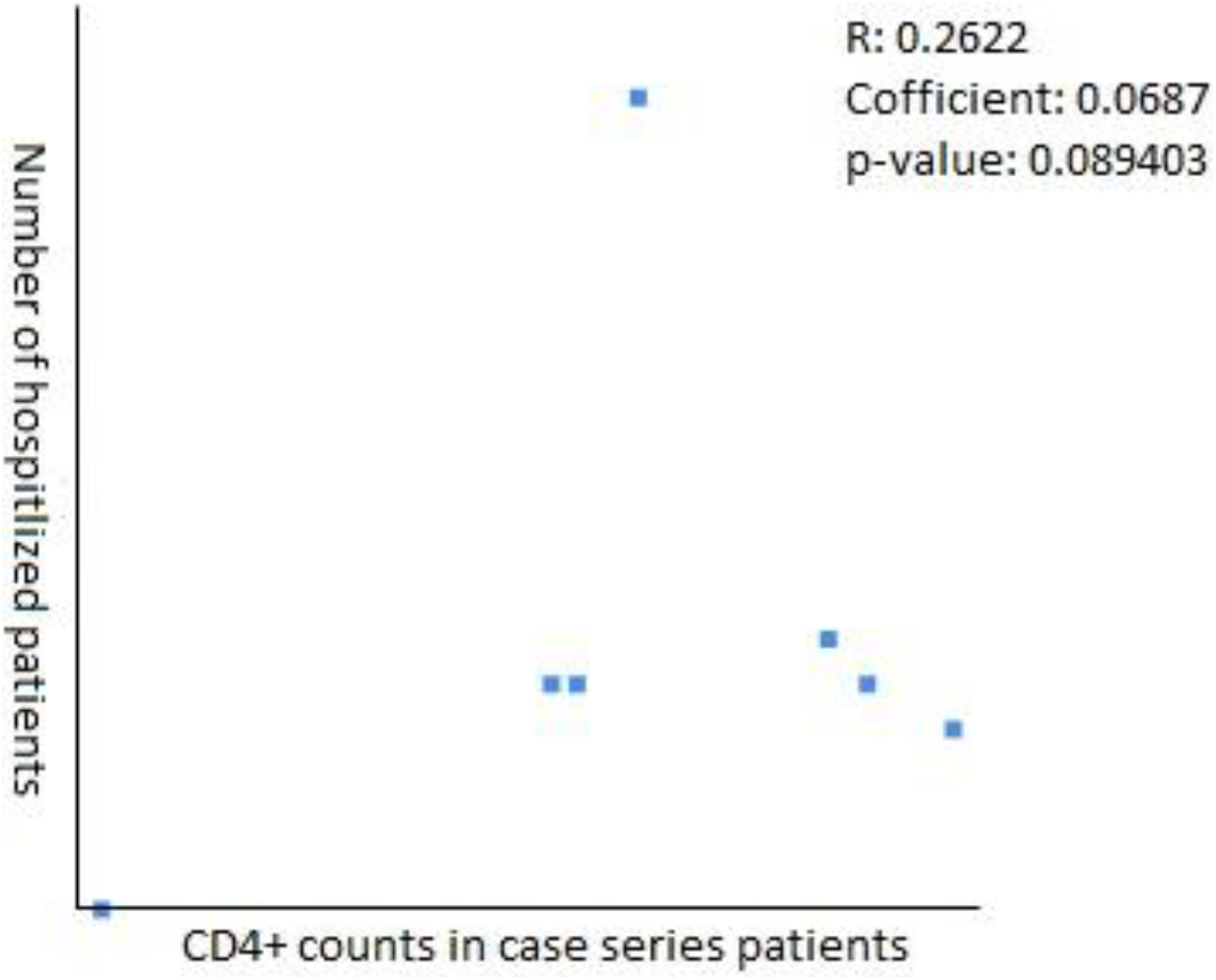
Correlation of CD4+ counts % hospitalization in case series

**Figure 3:**
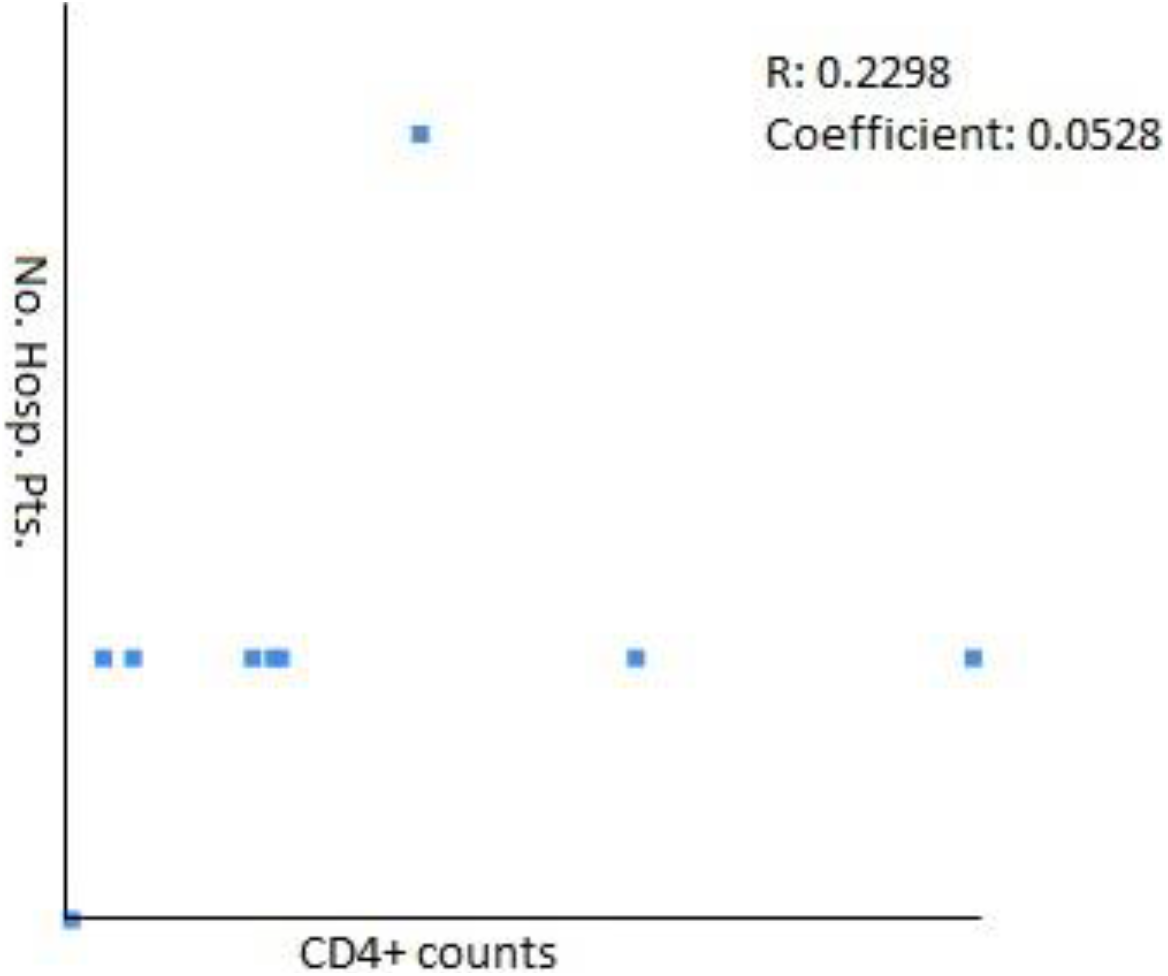
Correlation of CD4+ counts & hospitalization in Case Reports

**Figure 4.**
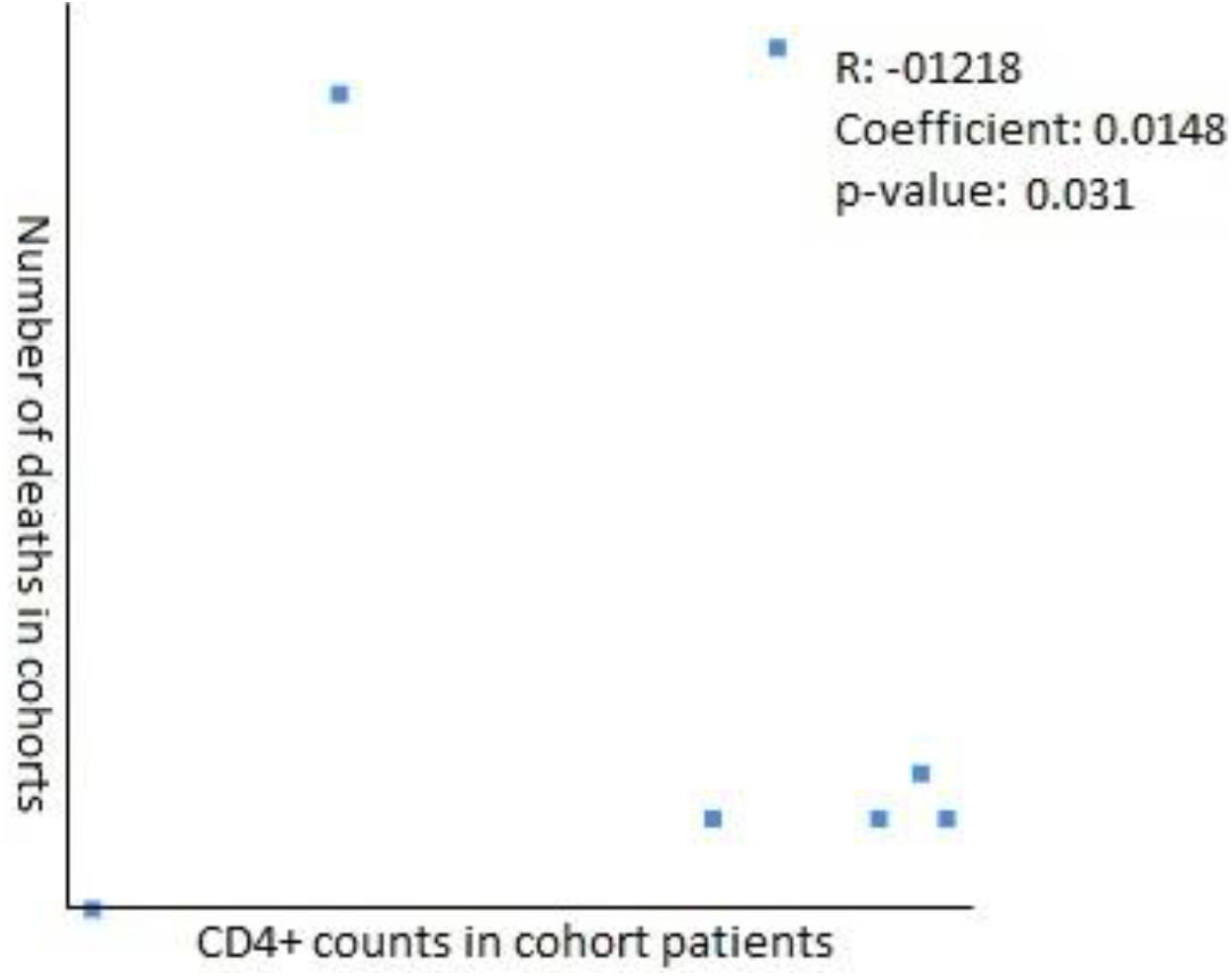
Correlation of CD4+ counts & Mortality in cohorts

**Figure 5.**
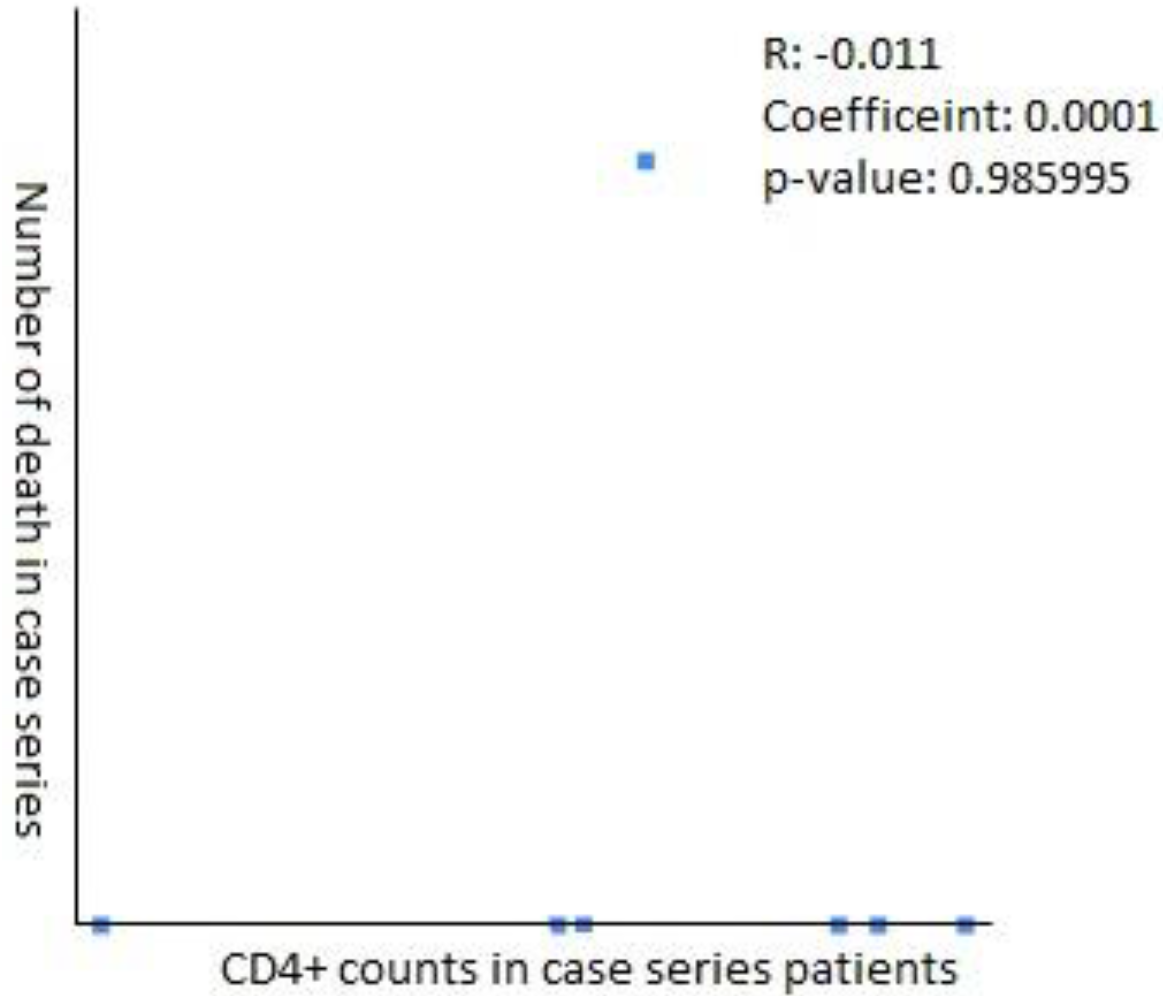
Correlation of CD4+ counts & Mortality in Case series

**Figure 6.**
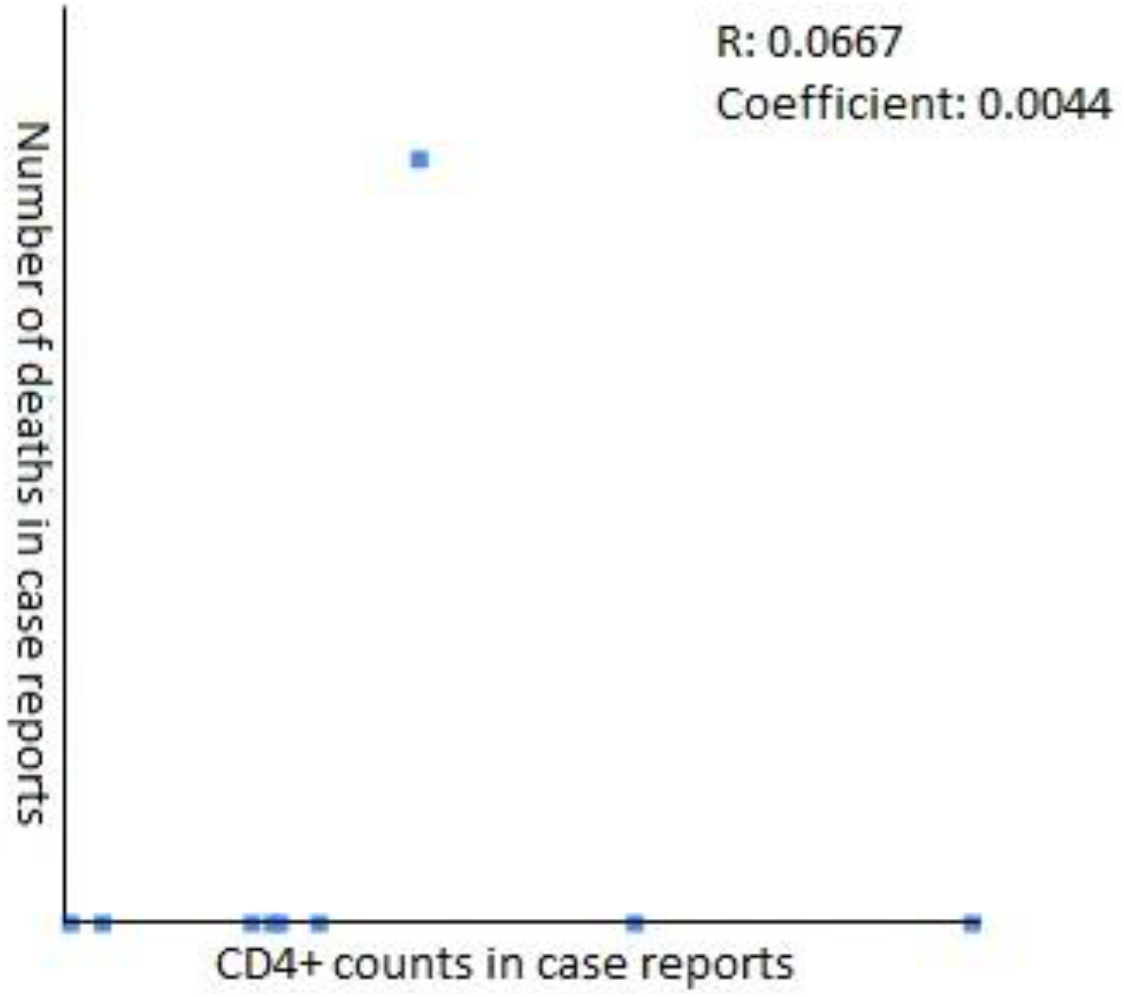
Correlation of CD4+ counts & Mortality in case reports

**Fig.7.**
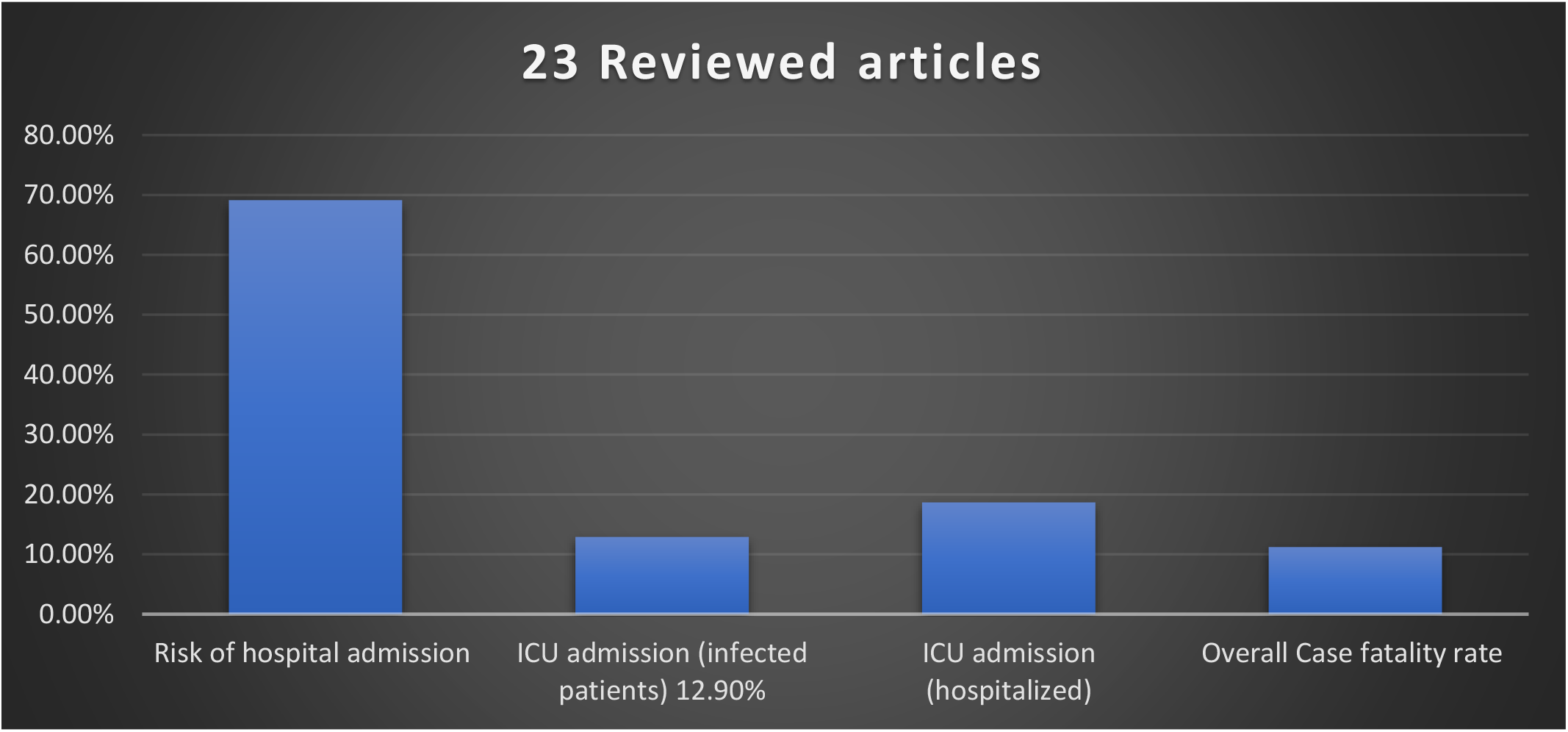
Results: As scoping review of HIV and COVID-19.

**Fig. 8.**
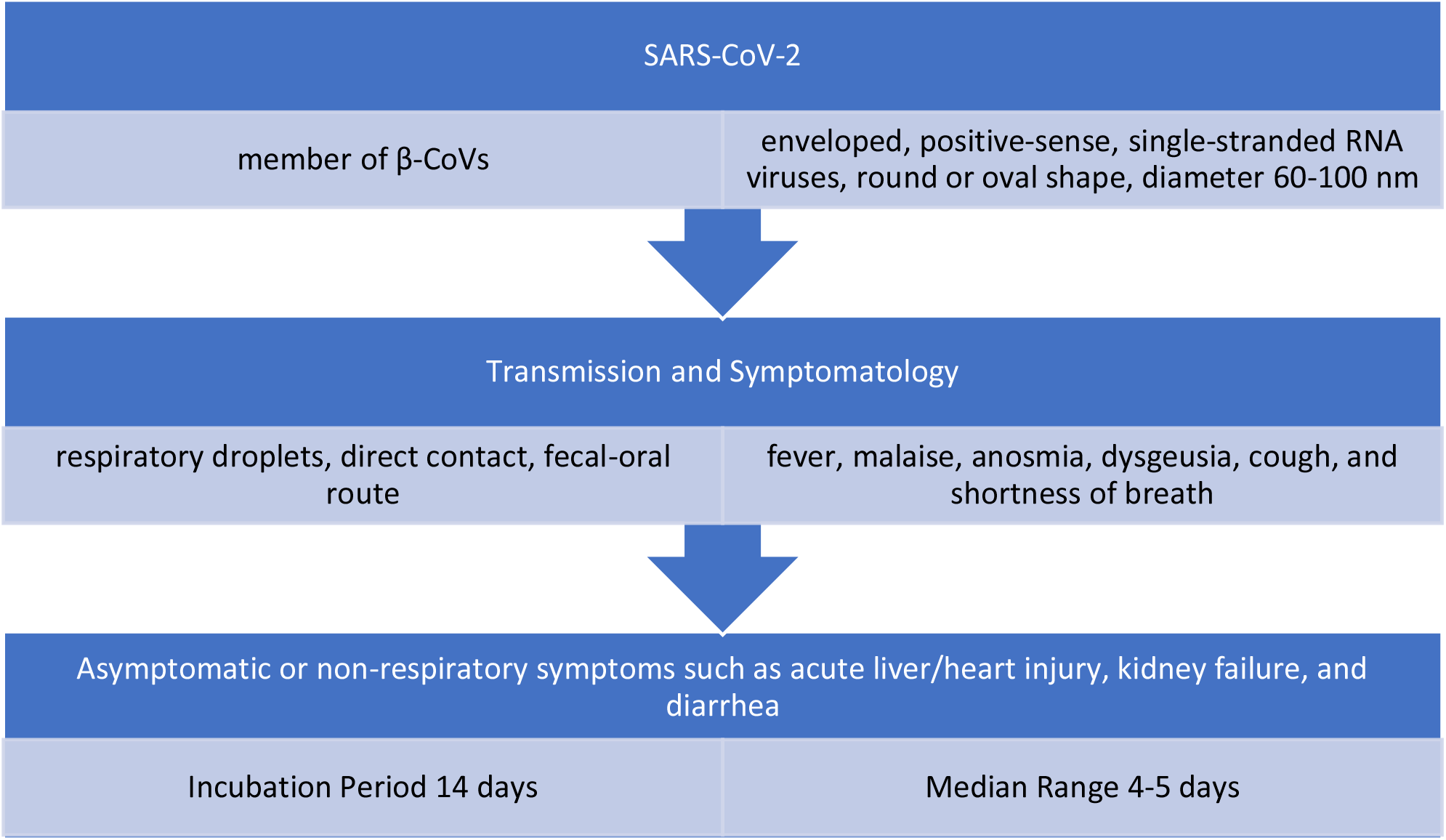
SARS-CoV-2 (Ref. 2- 10)

**Fig.9.**
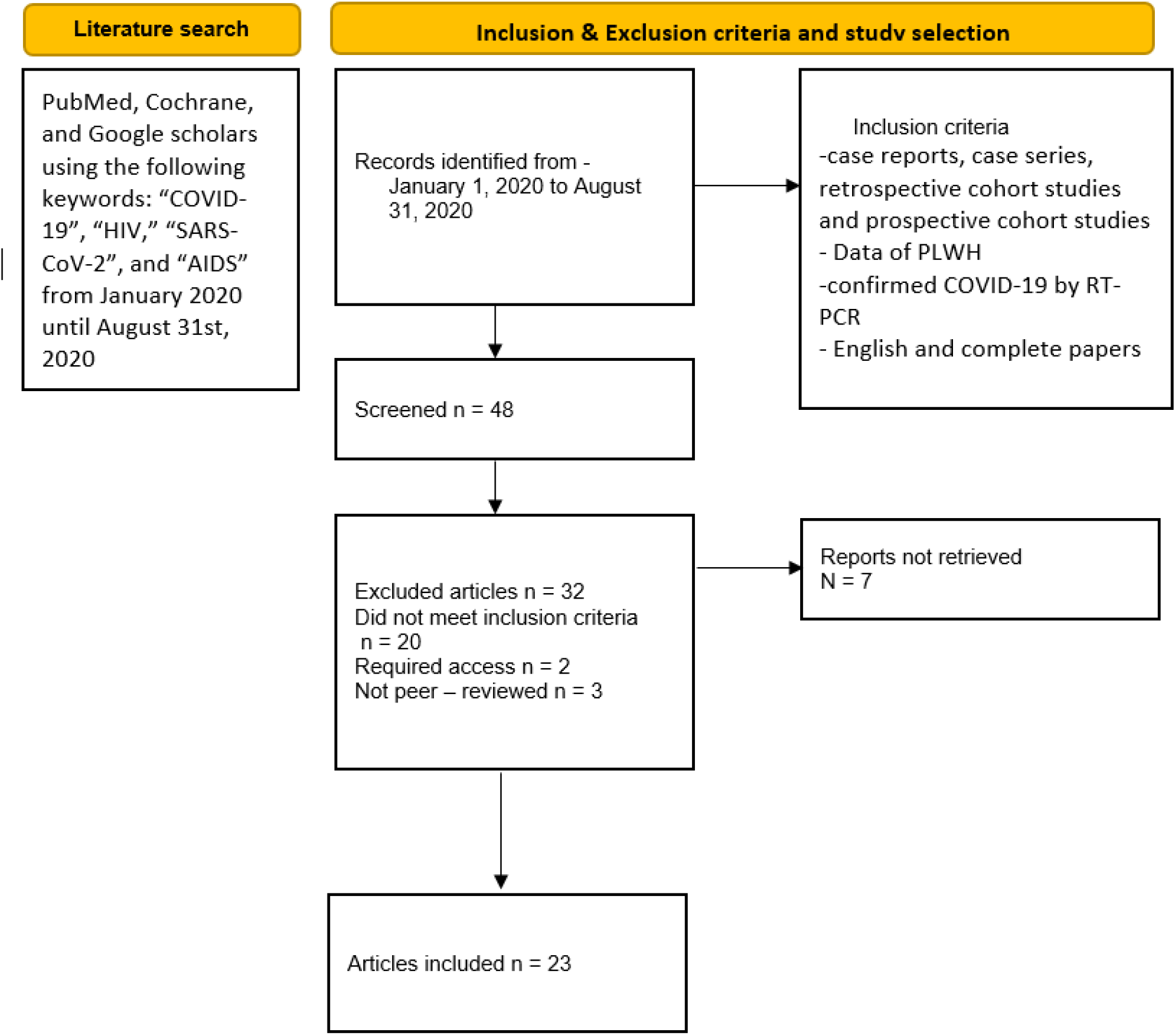
PRISMA guidelines for study acquisition.

**Fig. 10.**
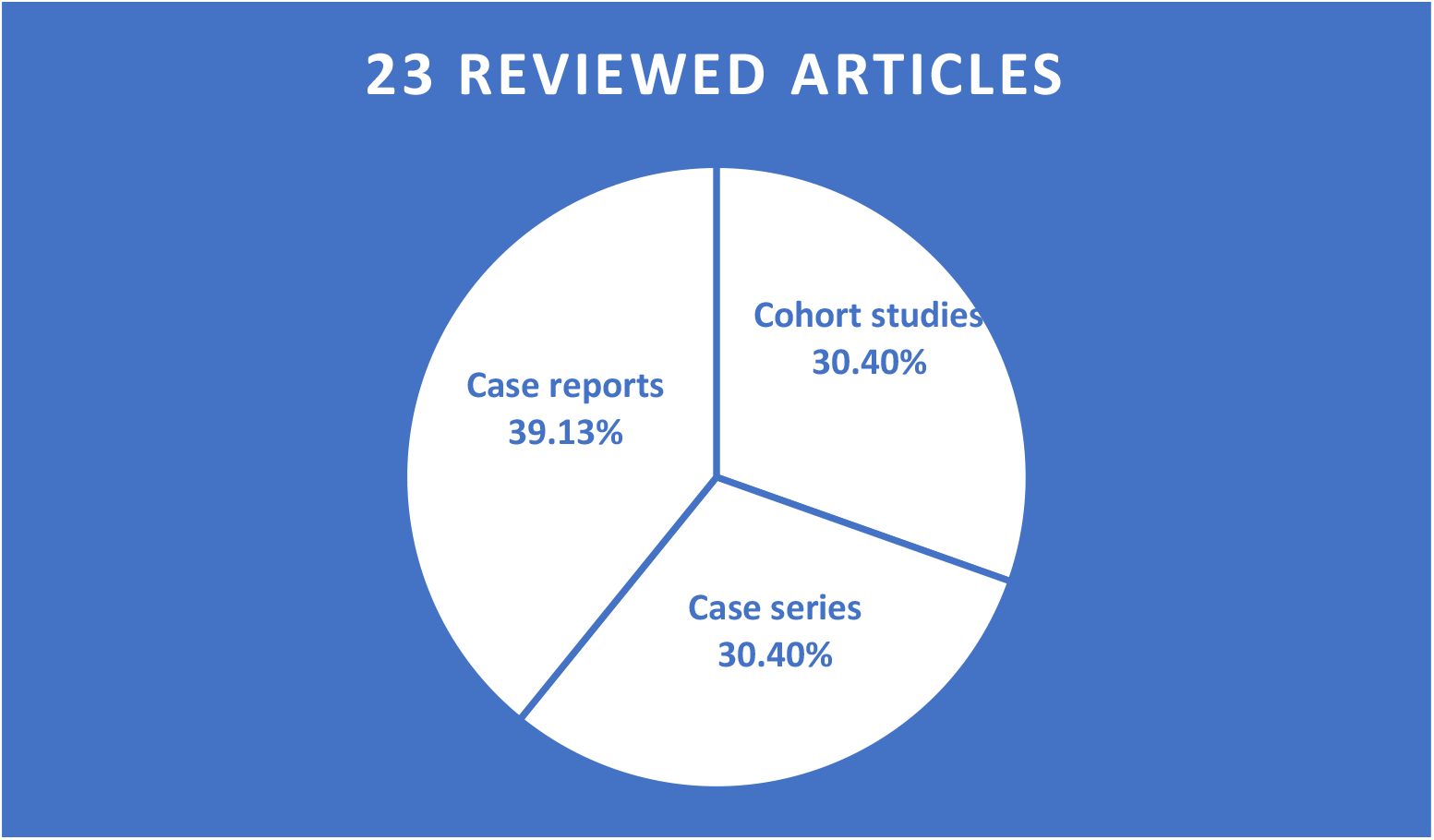
Types of included studies.

**Fig. 11.**
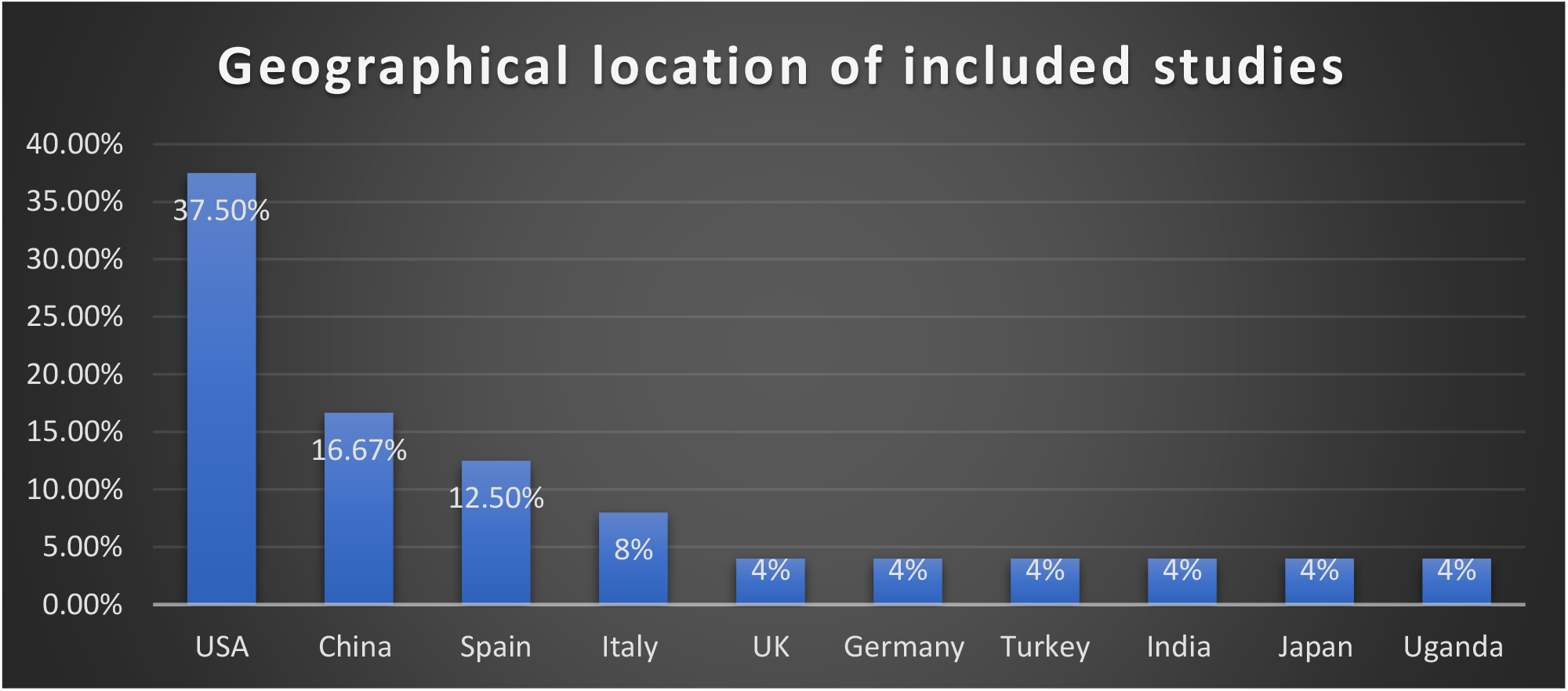
Geographical location of included studies.

### Clinical presentations in PLWH + COVID-19

The clinical presentation from the pooled data of the 23 articles was available for 319 patients. The most common symptoms among these patients were cough (57.4%, no= 183), fever (55.5%, no= 177), SOB (21.9%, no= 70), myalgia (13.2%, no= 42), fatigue (11.9%, no= 38), and headache/malaise (11%, no= 35). And least common with less than 10% were dyspnea (9.7%, no= 31), gastrointestinal symptoms (nausea/vomiting/diarrhea) (6.6%, no= 21), sore throat (6.3%, no= 20), anosmia/ageusia (4.4 %, no= 14), and altered mental status (4.4%, no= 14) [Table 5].

**Table 4.** Mortality data from all reviewed articles.

| Author (no. death) | Age (mean)<br>Gender | Comorbidities | ART |
| --- | --- | --- | --- |
| Viscarr et al (no= 2) | N/A | N/A | N/A |
| Del Amo et al (no= 20) | N/A<br>16 m, 4 f | N/A | -10 pt. on TAF/FTC<br>-8 pt. on ABC/3TC |
| Ho et al (no= 19) | 62 (55-68)<br>13 m, 6 f | 13 obesity | -10 pt. on TDF<br>-3 pt. on PI |
| Sigel et al (no= 18) | 62 (57-67)<br>13 m, 5 f | 4 DM, 6 HTN, 2 COPD, 6 CKD, 3 organ transplant, 1 liver cirrhosis, 1 obesity | -13 pt. on Integrase<br>-5 pt. on PI<br>-16 pt. on NRT |
| Meyerowitz et al (no= 2) | 59, 63<br>1 m, 1 f | 1st pt.: CHF, CKD, DM, HLD, HTN, prior stroke, 40 BMI<br>2nd pt.: HTN, prior stroke, CKD | -1st pt.: CTG+ABC/ETG+DRT/r+ETR<br>-2nd pt.: DTG+TAF/FTC |
| Gervasoni et al (no= 2) | 47, N/A<br>2 m | -1st pt.: obesity<br>-2nd pt.: CVD, lung cancer | N/A |
| **Harter et al (no= 3)** | 59, 55, 82<br>3 m | -1 pt.: HTN, COPD, DM<br>-other 2: N/A | -1st pt.: DOR/TDF/FTC<br>-2nd pt.: BIC/TAF/FTC<br>-3rd pt.: DRV/RTV/RGV |
| **Byrd et al (no= 1)** | 59<br>m | ESRD, cancer | EFV, ABC |

**Table 5.**
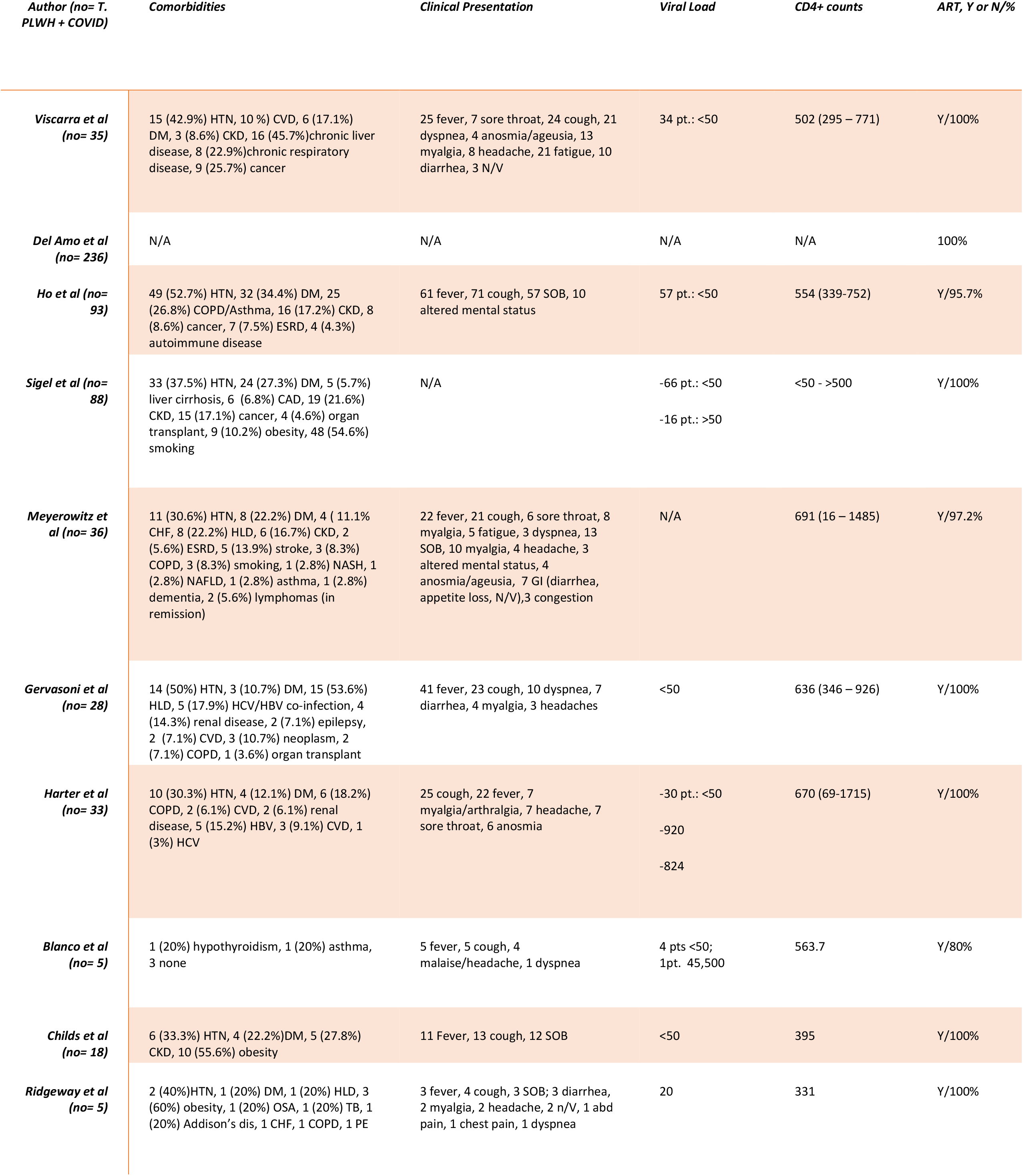
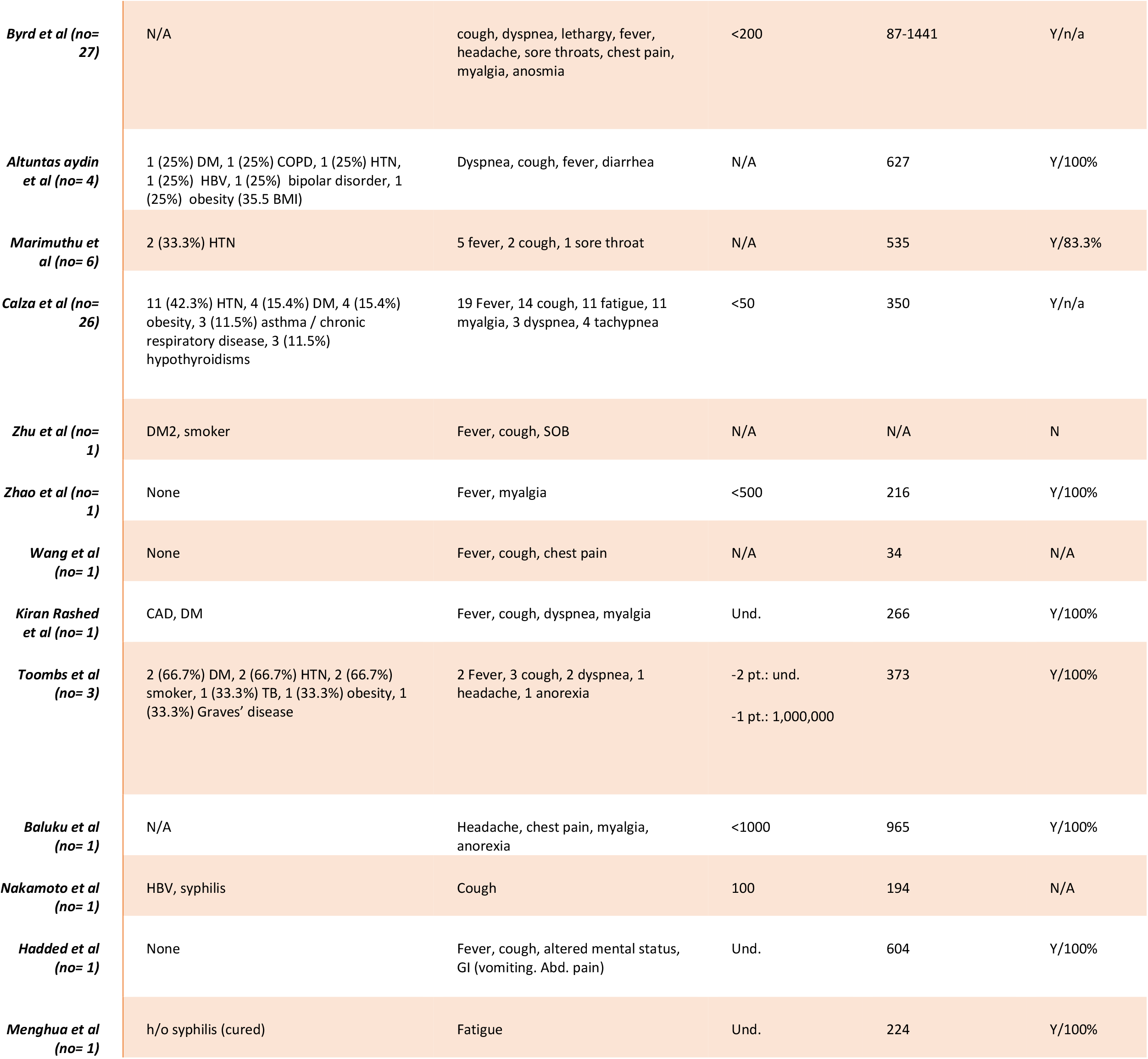
Patient Characteristics of all reviewed articles (comorbidity, CD4 counts, viral load)

### Co-morbidities in COVID positive PLWH groups

The comorbidities from the 23 articles were reported for 417 out of 649 patients; of which the most common chronic medical conditions found were hypertension (37.4%, no=156), diabetes mellitus (21.8%, no= 91), chronic kidney disease (12.2%, no= 51), COPD/asthma/lung disease (12.2%, no= 51), cancer (8.9%, no= 37), obesity (6.7%, no= 28), cardiovascular disease (6.2%,no= 26), chronic liver disease (5.5%, no= 23), and hyperlipidemia (5.8%, no= 24) [Table 5]

## Discussion

We conducted a systematic quantitative review to evaluate certain aspects of COVID-19 in patients living with human immunodeficiency virus (PLWH). The outcomes were categorized into risk of hospitalization, ICU admission, IMV requirement and mortality. The review of 23 articles indicated that there was a consistent high risk of hospitalization, ICU admission, IMV requirement and mortality in PLWH + confirmed COVID-19-positive in the cohort studies, case series and case reports. The rate of hospitalization from the pooled data of the 3 groups (Ch, CS, Cr) was 69.13% (n=450); 70.49% in Ch [12-18], 57.14% in CS [19-25], and 100% in CR [26-34]. The rate of ICU admission was 12.9% (n=84) for the 3 groups; 18.61% in Ch [12-18], 17.31% in CS [19-25], and 27.27% in CR [26-34]. And the overall case fatality rate of all reviewed articles was 11.21% (n=73).

Our results indicate that PLWH were a high risk group with greater than 50% needed hospitalization while 1/5 of hospitalized patients needed critical care. This finding aligns with the argument that predicts a bad outcome of COVID-19 in PLWH due to immunocompromised status [14, 20] and does not support the argument predicting a better prognosis for PLWH. [15, 17, 18, 19] The better prognosis could be explained by a lower risk for cytokine storm as a result of immunodeficiency, as there are low CD4 T lymphocytes which are responsible for activating innate immune cells like B-lymphocytes, cytotoxic T cells, and non-immune cells. There is also some suppression of the immune reaction through subtype T helper cells. [35]

Correlation analysis of CD4+ counts with hospitalization and mortality were not consistent in all reviewed studies as it showed weak positive in case series and case reports while weak negative in cohorts [12, 14-18]. And in mortality, there was a weak negative correlation in cohorts and case series [19-25] while weak positive in case reports [26-34]. More studies needed to focus on this to determine if CD4+ counts strongly correlate with the outcome of COVID-19.

We found that PLWH and confirmed COVID-19 do not present differently than the rest of the population as the common clinical presentations are cough (28.2%), fever (27.3%), and shortness of breath (10.8%), which is consistent with the finding of cohorts [12, 14, 16-18] and case series [19-25].

Analyzing the available data for deceased patients, we found the age range to be between 47 and 82 with a male to female ratio of 3:1, this correlates with the general population as data reveal that males had a mortality rate more than females. [37] The comorbidity data of deaths were available for 26 deceased individuals: HTN (no=9), DM (no=6), COPD (no=3), CKD (no=8), ESRD (no=1), CVD/CHF (no=2), organ transplant (no=1), prior stroke (no=2), cancer (no=2), liver cirrhosis (no=1), and obesity (no=3). The CD4+ counts ranged from below 50 to 800. The average CD4+ counts for Ho et al. [14], Meyerowitz et al. [16], and Harter et al. [18] of the deaths the cohort studies were 686, 426, and 389, respectively, and mortality rates in these cohorts were 20.5%, 5.6%, and 9.1%, respectively.

Nearly 98% of the cohort and 95% of the case series patients were being treated with ART at the time of diagnosis. According to Del Amo et al.[13], HIV-positive patients who were receiving TDF/FTC had a lower risk of contracting COVID-19 and ultimately being hospitalized, concluding that further investigation is necessary. However, Kate Child et al [20] concluded that patients on suppressive ART were not protected from moderate or severe COVID-19. We could not find any correlation between a specific type of ART and the risk of hospitalization, ICU admission, or mortality.

Despite the fact that this review was done following a standardized methodology, it presents with specific limitations. Cohort studies that we reviewed, such as Del Amo et al. and Meyerowitz et al. did not report an HIV viral load which makes it difficult to assess in relation to the other cohorts. Del Amo et al. also did not disclose a CD4+ count. Among the case reports, the HIV viral load and the total number of patients on IMV were not reported by Maomao et al. Lastly, Byrd et al. did not present the total number of patients admitted in the ICU and the number of patients on IMV. We did not run a study assessment because these limitations create barriers which make it hard to assess and compare the studies with one another.

## Conclusion

In conclusion, our results indicate that PLWH with COVID-19 are at increased risk for hospitalization, ICU admission, IMV requirement and mortality. We recommend classifying this group as high risk and they should adhere strictly to COVID-19 precautions. Authors conclude that larger sample size and study volume is needed to fully evaluate the outcome of this novel virus and confirm the association of CD4+cell counts with severe forms of the disease and mortality.

## Data Availability

A literature search was conducted with PubMed, Cochrane, and Google scholars using the following keywords: COVID-19, HIV, SARS-CoV-2, and AIDS from January 2020 until August 31st, 2020. Only papers in English language were considered. PRISMA guidelines were followed (Fig. 9)

## Acknowledgments

Krunal Pandav, MD; Sumedh Keul, statistician, Larkin Research Department, Miami, Florida

## Abbreviations

HIV: Human immunodeficiency virus
ART: Antiretroviral therapy
ICU: Intensive care unit
PLWH: People living with HIV
+ssRNA: positive single stranded RNA
SOB: shortness of breath
HTN: hypertension
DM: diabetes mellitus
CKD: chronic kidney disease
COPD: chronic obstructive pulmonary disease
CVD: cardiovascular disease
HLD: hyperlipidemia
HBV: hepatitis B virus
HCV: hepatitis C virus
CAD: coronary artery disease
ESRD: end stage renal disease
NASH: non-alcoholic steatohepatitis
OSA: obstructive sleep apnea
CHF: congestive heart failure
TB: tuberculosis

## Notes

### Competing Interest Statement

The authors have declared no competing interest.

### Funding Statement

There is no financial disclosure related to this study.

### Author Declarations

We did not seek IRB approval, as our study is based on data from published articles and patients were not directly involved.

